# HIV service interruptions during the COVID-19 pandemic in China: the role of HIV service challenges and institutional response from healthcare professional’s perspective

**DOI:** 10.1101/2021.03.12.21253463

**Authors:** Xueying Yang, Chengbo Zeng, Cheuk Chi Tam, Shan Qiao, Xiaoming Li, Zhiyong Shen, Yuejiao Zhou

**Affiliations:** South Carolina SmartState Center for Healthcare Quality, Arnold School of Public Health, University of South Carolina, Columbia, SC, USA; UofSC Big Data Health Science Center, University of South Carolina, Columbia, SC, USA; Department of Health Promotion, Education, and Behavior, Arnold School of Public Health, University of South Carolina, Columbia, SC, USA; Guangxi Center for Disease Control and Prevention, Nanning, Guangxi, China

**Keywords:** COVID-19, HIV service interruptions, HIV service challenges, Institutional response, China

## Abstract

**Background:** The healthcare system in China was largely overwhelmed during the unprecedented pandemic of coronavirus disease (COVID-19). HIV-related services have been unavoidably interrupted and impacted. However, the nature and scope of HIV service interruptions due to COVID-19 has rarely been characterized in China and how HIV service challenges affect the service interruptions are also unclear. The current study aimed at characterize HIV service interruption levels and analyzed its associated factors related to service challenges and institutional response from HIV healthcare providers’ viewpoint.

**Methods:** A cross-sectional online survey was conducted among 1,029 HIV healthcare providers in Guangxi, China, from April to May 2020. Latent class analysis (LCA) was first used to identify HIV service interruption levels. Then hierarchical multinomial logistic regression was conducted to analyze the relationships of HIV care service challenges and institutional response with HIV service interruption levels. Simple slope analysis was employed to examine interaction effects between HIV service challenges and institutional response to COVID-19.

**Results:** Four classes of HIV service interruption were identified using LCA, with 22.0% complete interruption (class 1), 15.4% moderate interruption (class 2), 21.9% minor interruption (class 3) and 40.7% almost no interruption (class 4). Using class 4 as a reference group, HIV care service challenges were positively associated with the probabilities of service interruptions (Class 1: AOR=1.23, 95%CI: 1.19∼1.26; Class 2: AOR= 1.10, 95%CI: 1.08∼1.13; Class 3: AOR= 1.10, 95%CI: 1.08∼1.12). Institutional response to HIV healthcare delivery was negatively associated with the probabilities of being classified into Class 1 (“Complete interruption”) (AOR=0.97, 95%CI: 0.93∼1.00) and Class 3 (“minor interruption [Outreach service]”) (AOR=0.96, 95%CI: 0.93∼0.99) as compared to Class 4 (“almost no interruption”). Institutional response to HIV healthcare delivery moderated the association of HIV service challenges with complete interruption, but not with the moderate or minor interruption when comparing with no interruption group.

**Conclusions:** A substantial HIV service interruptions occurs due to the COVID-19 pandemic, particularly services that require face-to-face interactions, such as VCT counselling, follow up and outreach services. HIV service challenges largely hinder the HIV service delivery. Institutional response to HIV healthcare delivery could marginally buffer the negative effect of service challenges on complete HIV service interruptions. To maintain continuity of core HIV services in face of a pandemic, build a resilient health care system with adequate preparedness is necessary.

## Introduction

The unprecedented pandemic of coronavirus disease (COVID-19) has not only triggered enormous human casualties and serious economic loss but also unbearable burdens to the health care system (1). Despite the strict containment measures implemented in China had reduced transmission, the healthcare system was largely overwhelmed by such a public health emergency (2). HIV-related services have been unavoidably interrupted and impacted. The U.S. Center of Disease Control and Prevention (CDC) posted specific guideline to address concerns and questions of people living with HIV (PLWH) related to their COVID-19 risk and prevention(3). Chinese government has issued guidelines related to how to deal with the dual pandemic of HIV and COVID-19. A survey of programmes supported by the Global Fund to Fight HIV, Tuberculosis, and Malaria, in 106 countries showed that disruptions to service delivery due to the COVID-19 pandemic have affected 85% of HIV programmes (4).

The COVID-19 pandemic has posed various challenges to maintaining HIV care. First, many public health organizations (especially hospitals) put COVID-19 control in first priority. Numerous HIV healthcare providers were reassigned to fight against the COVID-19 outbreak given the shared logistics with infectious disease facilities. As a results, various HIV service delivery were put aside due to the personnel shortage and the ongoing needs of PLWH could not be met (5). Second, the emergent pandemic prevention measures, such as stringent quarantine enforcement and transportation lockdown in various cities across China has also severely impeded both HIV patients and healthcare providers to seek or provide HIV services (5). Third, the HIV clinics may lack personal protective equipment, and encounter no safe options/passage for transportation to and from work. Some staffs may be asked to task-shift or fill in for others who are sick or at high-risk for severe COVID-19. All these challenges have posed great threat to the HIV service delivery.

Consequently, these service challenges has led to various service interruptions in HIV clinics, and the level of interruptions varies across countries (6). First, the routine, non-urgent health care visits and supportive services which requires face-to-face counselling are unavoidably affected due to the strict lockdown. For example, a US study suggested that nearly 56% of the HIV clinics were partially interrupted and 26% were completely closed (7). According to a survey conducted in 19 European countries, only six countries operated normally; 11 countries’ physicians were sharing HIV and COVID-19 care duties, although no country reported complete HIV clinic closures(8). In China, a national anonymous survey conducted by Joint United Nations Programme on HIV/AIDS (UNAIDS) and China CDC reported that 32.6% of Chinese PLWH were at risk of antiretroviral therapy (ART) discontinuation during the pandemic and 48.6% did not know where to acquire antiretroviral drugs in the near future (9, 10). Moreover, high-risk populations cannot get access to pre-exposure and post-exposure prophylaxis (PrEP and PEP) services on time, which might result in resurgence of new cases of HIV infection (11).

Disruption to delivery of HIV health care caused by COVID-19 could lead to adverse consequences for individuals’ health beyond those from COVID-19 itself (12-14). For instance, prediction models from sub-Saharan Africa found that a 6-month interruption of antiretroviral drug supply across 50% of the population of PLWH who are on ART could lead to a 1.63 times increase in HIV-related deaths over a 1-year period compared with no disruption(15). In addition, interruption of ART would increase mother-to-child transmission of HIV by approximately 1.6 times(15). WHO and UNAIDS literature has also highlighted severe consequences of disruptions, stating that a 6-month disruption of ART could lead to more than 500,000 extra deaths from AIDS-related illnesses(16). Therefore, a thorough investigation in terms of the depth and breadth of HIV service interruption (e.g., levels of interruption and particular services that were disrupted) are warranted. In addition, factors that influence the impact of HIV service challenges on different levels of HIV service interruption are also unclear.

Institutional responses to HIV health care delivery in the context of COVID-19 pandemic could possibly buffer the negative effect of HIV service challenges on HIV service interruption. Institutional responses in our context refers to strategies or measures that implemented at organizational level to help maintain the regular HIV service provision in the amidst of COVID-19 pandemic. In some countries, the COVID-19 pandemic and associated national lockdowns have led to increased innovations and ingenuity in HIV service delivery. For example, to ensure the continuity of ART supply, South Africa have implemented extended refills of ART, support out-of-facility pickup points for refills, transition counselling and adherence support to virtual platforms, coordinate the local NGOs for ART distribution (17-20). These institutional responses might play an essential role in building a functional and resilient health systems, but scant literature on COVID-19 has examined their effects from a HIV-service-delivery perspective. What institutional responses have been implemented in Chinese HIV clinics and whether these strategies respond effectively to the HIV service interruptions are unclear. Thus, it is worth investigating institutional response and its role in buffering HIV service challenge influences on different levels of HIV service interruptions during the COVID-19 pandemic.

In sum, in order to improve healthcare delivery and provide sustainable quality care to PLWH during the COVID-19 pandemic and future pandemics, we need a better understanding of the scope of HIV treatment service interruptions and the roles of HIV service challenges and institutional response to HIV healthcare delivery in exacerbating or alleviating such service interruptions. Specifically, this paper aims to explore the levels of HIV service interruptions in China and examine the main and interactive effects of HIV service challenges and institutional response on the levels of HIV service interruptions.

## Methods

### Participants and data collection

Data collection of the current cross-sectional study was conducted using an anonymous online survey from a convenience sample of HIV healthcare providers in Guangxi Zhuang Autonomous Region (“Guangxi”) from April through May 2020. Guangxi, a region located in Southern China, was consistently ranked third in terms of the number of HIV infection cases across the 31 provinces from 2014 to 2018 (21). The study design and data collection procedure have been described in detailed elsewhere (22). The inclusion criteria for participants included: 1) currently a healthcare provider offering HIV-related care and services; 2) 18 years of age or older; and 3) living in Guangxi. First, staff in Guangxi CDC contacted HIV healthcare providers by email. Once the healthcare providers’ eligibility was determined, local staff would invite them to participate in the online survey. Participants were also encouraged to share the invitation letter with their colleagues. An electronic consent was provided prior to the survey started. A total of 1,280 HIV healthcare providers consented and completed the survey. Among them, 251 participants were excluded as they were identified as someone from outside of Guangxi (*n*=63), or their responses were considered as random or careless answers (*n*=76) or outliers on multiple questions (*n*=112), resulting in a final sample size of 1,029 in this study. The research protocol was approved by the Institutional Review Boards at both the University of South Carolina in the United States and the Guangxi CDC in China.

### Measures

#### Institute levels

Healthcare providers were asked to report their institute levels, which were classified into province-, city-, county-, and rural-levels. Due to the small sample sizes (16.6%) in both province- and city-levels, we dichotomized it into province/city level and county/rural level.

#### HIV care service interruptions

Healthcare providers were asked whether there were any interruptions in HIV care services (i.e., HIV clinical service, voluntary counselling and testing [VCT] service, treatment initiation service, outreach work, follow-up service, and ART provision) in their institutes due to COVID-19 using a six-item self-developed checklist (Table 1). The response options for each type of HIV care service interruptions were “Yes (1)” and “No (0)”. This checklist had good reliability among the study sample (Cronbach’s alpha=.87).

**Table 1.** Proportion of different HIV service interruptions (*n*=1,029)

| Items | Yes ($n$, %) |
| --- | --- |
| HIV clinic service suspended because of COVID-19 | 52 (5.05) |
| The VCT service is suspended or postponed because of the COVID-19 | 555 (53.94) |
| The ART application service is suspended or postponed | 692 (67.25) |
| The outreach work is suspended or postponed | 351 (34.11) |
| The follow up service is suspended or postponed | 76 (7.39) |
| Suspended or postponed ART provision due to the short supply | 5 (0.49) |

#### HIV service challenges

HIV service challenges in participants’ institutes were assessed using a self-developed scale comprised of eight items about perceived difficulties and barriers of delivering HIV services during the COVID-19 pandemic. Sample items included “Could not get to work because of self-quarantine or traffic restrictions” and “No manual or guidelines”. Response option was rated from “Strongly disagree (1)” to “Strongly agree (5)”. A sum score was calculated with higher score indicating a higher level of HIV service challenges. This scale showed good reliability in the current study sample (Cronbach’s alpha=.87).

#### Institutional response to COVID-19

An 11-item self-developed measure was employed to assess institutional response to the COVID-19 pandemic, such as measures to prevent COVID-19 transmission, training and guidance. Given that our outcomes are HIV service interruption, we used the 5-item subscale ‘healthcare delivery’ to reflect the institutional response to HIV service provision during COVID-19 pandemic. Sample items were “Dispensed antiretroviral medications for 3-6 months at once to ensure ample medications for HIV patients” and “Posted online information about HIV care services”. Response option was ranging from “Never (1)” to “Always (5)”. A sum score was calculated with a higher score representing a higher level of utilizing these responses in the response to COVID-19 pandemic. The internal consistency of this scale was adequate (Cronbach’s alpha=.72).

### Statistical analysis

Data analyses included in this study were latent class analysis (LCA), descriptive statistics, bivariate analysis, and hierarchical multinomial logistic regression. First, LCA was used to identify the interruption levels using the six-item checklist. Based on the standard procedure of mixture modelling (23), a baseline, single-group model was specified. Then, successive models with increasing numbers of subgroup were fitted. The final number of subgroups or final model was determined based on the model interpretation, size of estimated subgroup proportions and model fit indices including Log Likelihood values, Akaike Information Criterion (AIC), Bayesian Information Criterion (BIC), adjusted BIC, entropy, *p*-value of bootstrap likelihood ratio test (BLRT). The interruption level was interpreted based on the item-response probabilities by each subgroup and assessed in accordance with prior research. Estimated subgroups with less than 5% of the total sample were not considered due to the possibility of class over-extraction and poor generalizability (24). Lower absolute values on the information criterion (i.e., AIC, BIC, adjusted BIC) indicate better fitting models(23, 25). The final model was identified when entropy was larger than 0.80 and the *p*-value of BLRT were insignificant.

Second, descriptive statistics were reported on institute levels, HIV service challenges, and institutional response to HIV healthcare delivery by the identified interruption levels. Median and interquartile range (IQR) were used to describe continuous variables while frequency and percentage were used for categorical variable. Third, Chi-square test or analysis of variance (ANOVA) were used to examine the bivariate relationships between variables of interest and interruption levels.

Finally, hierarchical multinomial logistic regression was conducted to predict interruption levels using institute levels, HIV service challenges, and institutional response to HIV healthcare delivery during COVID-19 pandemic. First, institute levels were adjusted in model 1. HIV service challenges and institutional response to HIV healthcare delivery were entered to model 2 and evaluated their impacts on interruption levels after accounting for institute levels. Then, an interaction term between HIV service challenges and institutional response was created and entered to model 3. To avoid collinearity, variables were centralized before the creation of interaction. Simple slope analysis was used to examine the interactions. LCA was conducting using Mplus version 7.0 (Muthen & Muthen, Los Angeles, CA) while descriptive statistics, bivariate analysis, and hierarchical multinomial logistic regression were conducted using SAS software version 9.4 (SAS Institute, Inc., Cary, NC).

## Results

### Latent class analysis

The absolute values of Log Likelihood, AIC, and adjusted BIC decreased with the increasing numbers of subgroups while BIC increased at the 5-class solution as compared to 4-class solution. Additionally, the smallest class at the 5-class solution included less than 5.0% of the sample size. Accordingly, the 4-class solution was selected as it accounted for a better classification. Table 2 shows the model fits of each solution in LCA.

**Table 2.** Model fits (*n*=1,029)

| Models | *LL* | *AIC* | *BIC* | *aBIC* | *Entropy* | *BLRT* | *% of the smallest class* |
| --- | --- | --- | --- | --- | --- | --- | --- |
| Baseline | -3976.01 | 7964.02 | 7993.64 | 7974.58 | - | - | - |
| 2 | -2910.06 | 5846.11 | 5910.29 | 5869.0 | 0.88 | <0.001 | 39.98 |
| 3 | -2744.73 | 5529.46 | 5628.18 | 5564.66 | 0.85 | <0.001 | 25.22 |
| **4** | **-2695.01** | **5444.02** | **5577.31** | **5491.55** | **0.86** | **<0.001** | **16.56** |
| 5 | -2670.84 | 5409.67 | 5577.51 | 5469.52 | 0.91 | <0.001 | 4.97 |
| 6 | -2658.79 | 5399.57 | 5601.96 | 5471.74 | 0.90 | 0.013 | 1.58 |

*Note:* *LL*: Log likelihood value.

Table 3 presents the probability of class memberships. Across the four classes, most HIV service interruption items in Class 4 had the lowest probability compared with other classes and was named as “*Almost no interruption (420 [40*.*8%])*”. In contrast, items in Class 1 had the highest probability and was named as “*Complete interruption (226 [22*.*0%])*”. Several items in Class 2 had high probabilities such as HIV clinic service, VCT service, outreach service and follow-up service and was named as “*Moderate interruption (VCT + treatment initiation service + Outreach service) (158 [15*.*4%])*”. Only two items (outreach service and follow up service) in Class 3 had high probabilities and was then named as “*Minor interruption (Outreach service) (225 [21*.*9%])*”.

**Table 3.** Probability of class membership

| Items | Class 1 | Class 2 | Class 3 | Class 4 |
| --- | --- | --- | --- | --- |
| Sample sizes ($n$, %) | 226 (22.0) | 158 (15.4) | 225 (21.9) | 420 (40.7) |
| HIV clinic service suspended because of COVID-19 | **0.931** | **0.743** | 0.000 | 0.021 |
| The VCT service is suspended or postponed because of the COVID-19 | **1.000** | **0.834** | 0.197 | 0.030 |
| The ART application service is suspended or postponed | **0.998** | 0.249 | 0.141 | 0.012 |
| The outreach work is suspended or postponed | **0.998** | **0.896** | **0.968** | 0.311 |
| The follow up service is suspended or postponed | **0.982** | **0.754** | **0.970** | 0.038 |
| Suspended or postponed ART provision due to the short supply | **0.961** | 0.291 | 0.279 | 0.040 |

*Note:*
*Class 1: Completed interruption*
*Class 2: Moderated interruption (VCT + Treatment initiation + Outreach service)*
*Class 3: Minor interruption (Outreach service)*
*Class 4: Almost no interruption*

### Descriptive statistics and bivariate analysis

Among 1,029 healthcare providers, 859 (83.5%) worked in the county/rural level-institutes. The *medians* and *IQR*s of HIV service challenges and institutional response to HIV healthcare delivery were 25.0 (18.0∼32.0) and 17.0 (13.0∼21.0), respectively.

Chi-square test suggested that institute affiliation levels were significantly associated with interruption levels (*p*=0.003). Particularly, 205 (90.7%) of healthcare providers reported *complete interruption* were from county/rural institutes. HIV service challenges was also significantly related to interruption levels (*p*<0.001). Participants who reported complete interruption perceived that their institutes had more HIV service challenges than those who did not. Results from ANOVA suggested that the level of institutional response to HIV healthcare delivery was related to interruption levels (*p*=0.002). Healthcare providers who reported *almost no interruption* perceived a higher level of institutional response in their institute than those who reported somewhat interruptions. Table 4 presents the results of descriptive statistics and bivariate analysis.

**Table 4.** Descriptive statistics and bivariate analysis

|  | Total | Class 1 | Class 2 | Class 3 | Class 4 | P-value |
| --- | --- | --- | --- | --- | --- | --- |
| **Institute types** |  |  |  |  |  | **0.003** |
| Province/city | 170 (16.5) | 21 (9.3) | 22 (13.9) | 46 (20.4) | 81 (19.3) |  |
| County/rural | 859 (83.5) | 205 (90.7) | 136 (86.1) | 179 (79.6) | 339 (80.7) |  |
| **Institutional response (Median, IQR)** | 17.0 (13.0, 21.0) | 17.0 (13.0, 21.0) | 17.0 (13.0, 21.0) | 16.0 (12.0, 21.0) | 18.0 (15.0, 21.0) | **0.002** |
| **HIV service challenges (Median, IQR)** | 25.0 (18.0, 32.0) | 32.0 (28.0, 38.0) | 27.0 (22.0, 32.0) | 26.0 (21.0, 32.0) | 19.0 (12.0, 26.0) | **<0.001** |

*Note: IQR: Interquartile range.*
*Class 1: Completed interruption*
*Class 2: Moderated interruption (VCT + Treatment initiation + Outreach service)*
*Class 4: Almost no interruption*

### Hierarchical multinomial logistic regression

Using “*Almost no interruption*” as a comparison group, hierarchical multinomial logistic regression was conducted. Results of model 1 suggested that healthcare providers from county/rural-level institutes had a higher probability to be classified into Class 1 (“*Complete interruption*”) as compared to those from province/city-level institutes (*AOR*=2.33, 95%*CI*: 1.40∼3.89). Model 2 found that HIV service challenges were significantly and positively associated with the probabilities of service interruptions (Class 1: *AOR*=1.23, 95%*CI*: 1.19∼1.26; Class 2: *AOR*= 1.10, 95%*CI*: 1.08∼1.13; Class 3: *AOR*= 1.10, 95%*CI*: 1.08∼1.12). Institutional response to HIV healthcare delivery could decrease the probabilities of being classified into Class 1 (“*Complete interruption”*) (*AOR*=0.97, 95%*CI*: 0.93∼1.00, p=0.048) and Class 3 (“*Minor interruption [Outreach service]*”) (*AOR*=0.96, 95%*CI*: 0.93∼0.99) as compared to Class 4 (“*Almost no interruption*”).

Finally, in model 3, significant interaction between HIV service challenges and institutional response could be found in the comparison between complete interruption and no interruption groups (*AOR*=0.72, 95%*CI*=0.56∼0.92). Under the same HIV care service challenges, healthcare providers from the institutes with more usage of institutional response to HIV healthcare delivery during COVID-19 pandemic were less likely to be classified into Class 1 (“*Complete interruption*”). Similar with models 1 and 2, relationships between institute levels, institutional response to HIV healthcare delivery during COVID-19, and interruption levels could also be founded in model 3. Table 5 shows the results of hierarchical multinomial logistic regression. Figures 1, 2, and 3 are the results of simple slope analyses.

**Table 5.** Multinomial logistics regression

| Variables | Class 1 vs. 4 |  | Class 2 vs. 4 |  | Class 3 vs. 4 |  |
| --- | --- | --- | --- | --- | --- | --- |
|  | AOR | 95%CI | AOR | 95%CI | AOR | 95%CI |
| **Model 1:** |  |  |  |  |  |  |
| **Institute** |  |  |  |  |  |  |
| County/rural | **2.33\*\*** | **1.40~3.89** | 1.48 | 0.89~2.46 | 0.93 | 0.62~1.39 |
| Province/city | Reference |  |  |  |  |  |
| **Model 2:** |  |  |  |  |  |  |
| **Institute** |  |  |  |  |  |  |
| County/rural | **2.23\*\*** | **1.26~3.96** | 1.46 | 0.86~2.49 | 0.93 | 0.60~1.43 |
| Province/city | Reference |  |  |  |  |  |
| **Institutional response** | **0.97\*** | **0.93~1.00** | 0.98 | 0.94~1.01 | **0.96\*\*** | **0.93~0.99** |
| **HIV service challenges** | **1.23\*\*\*** | **1.19~1.26** | **1.10\*\*\*** | **1.08~1.13** | **1.10\*\*\*** | **1.08~1.12** |
| **Model 3:** |  |  |  |  |  |  |
| **Institute** |  |  |  |  |  |  |
| County/rural | **2.24\*\*** | **1.26~3.99** | 1.47 | 0.86~2.51 | 0.93 | 0.61~1.43 |
| Province/city | Reference |  |  |  |  |  |
| **Institutional response (I)** | 0.93 | 0.74~1.17 | 0.84 | 0.69~1.03 | **0.75\*\*** | **0.63~0.90** |
| **HIV service challenges (C)** | **6.53\*\*\*** | **5.03~8.49** | **2.52\*\*\*** | **2.01~3.16** | **2.41\*\*\*** | **1.97~2.94** |
| **I\*C** | **0.72\*** | **0.56~0.92** | 0.85 | 0.68~1.06 | 0.93 | 0.77~1.13 |

*Note:* AOR: Adjusted odds ratio; CI: Confidence interval; \*: $p < 0.05$ ; \*\*: $p < 0.01$ ; \*\*\*: $p < 0.001$ .
*Class 1: Completed interruption*
*Class 2: Moderated interruption (VCT + Treatment initiation + Outreach service)*
*Class 3: Minor interruption (Outreach service)*
*Class 4: Almost no interruption*

**Figure 1.**
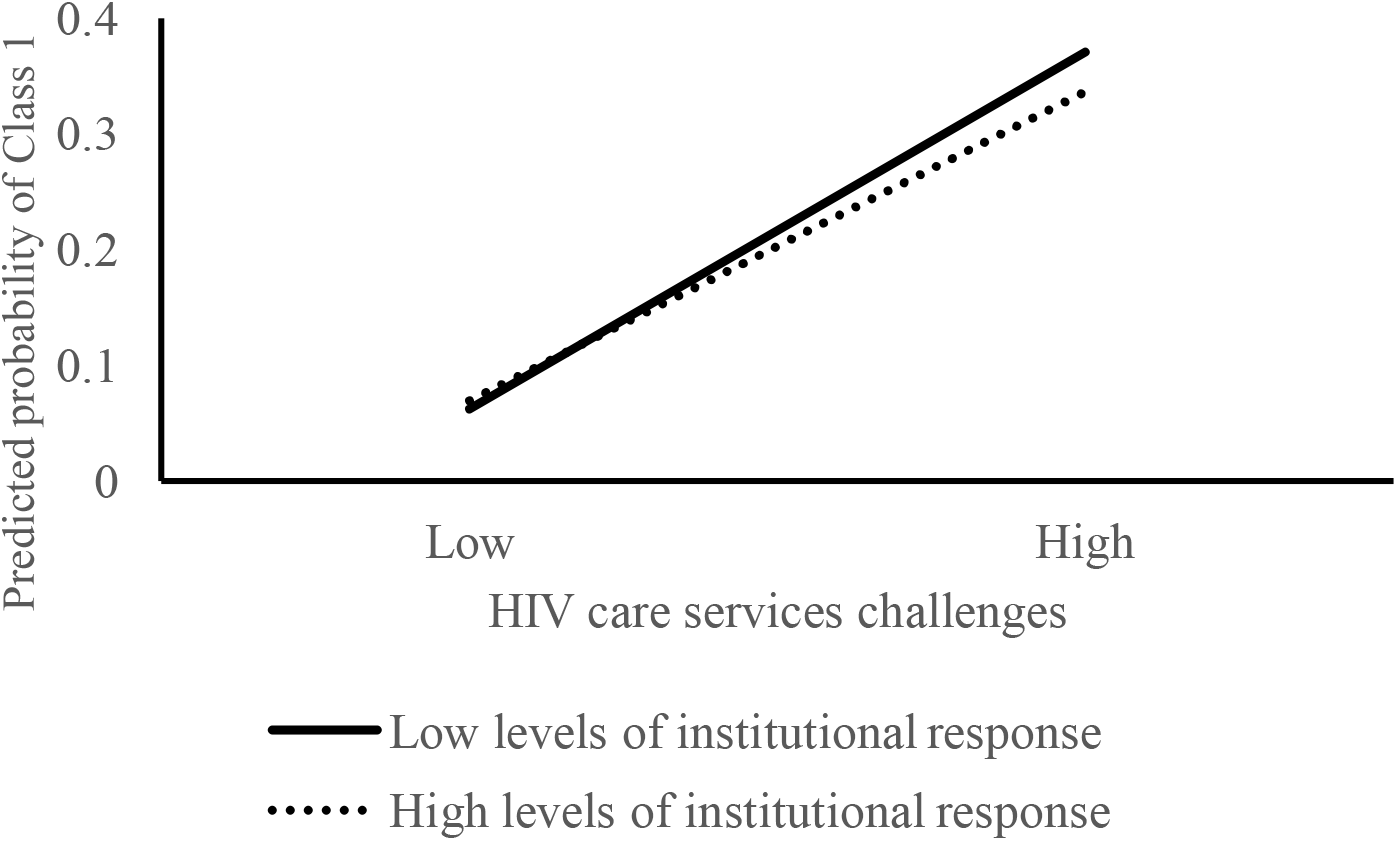
Interaction between HIV care services challenges and institutional response in the comparison between Class 1 (Completed interruption) and Class 4 (Minor interruption) ***Note:*** *Higher and lower are defined as values above or below the mean, respectively*.

**Figure 2.**
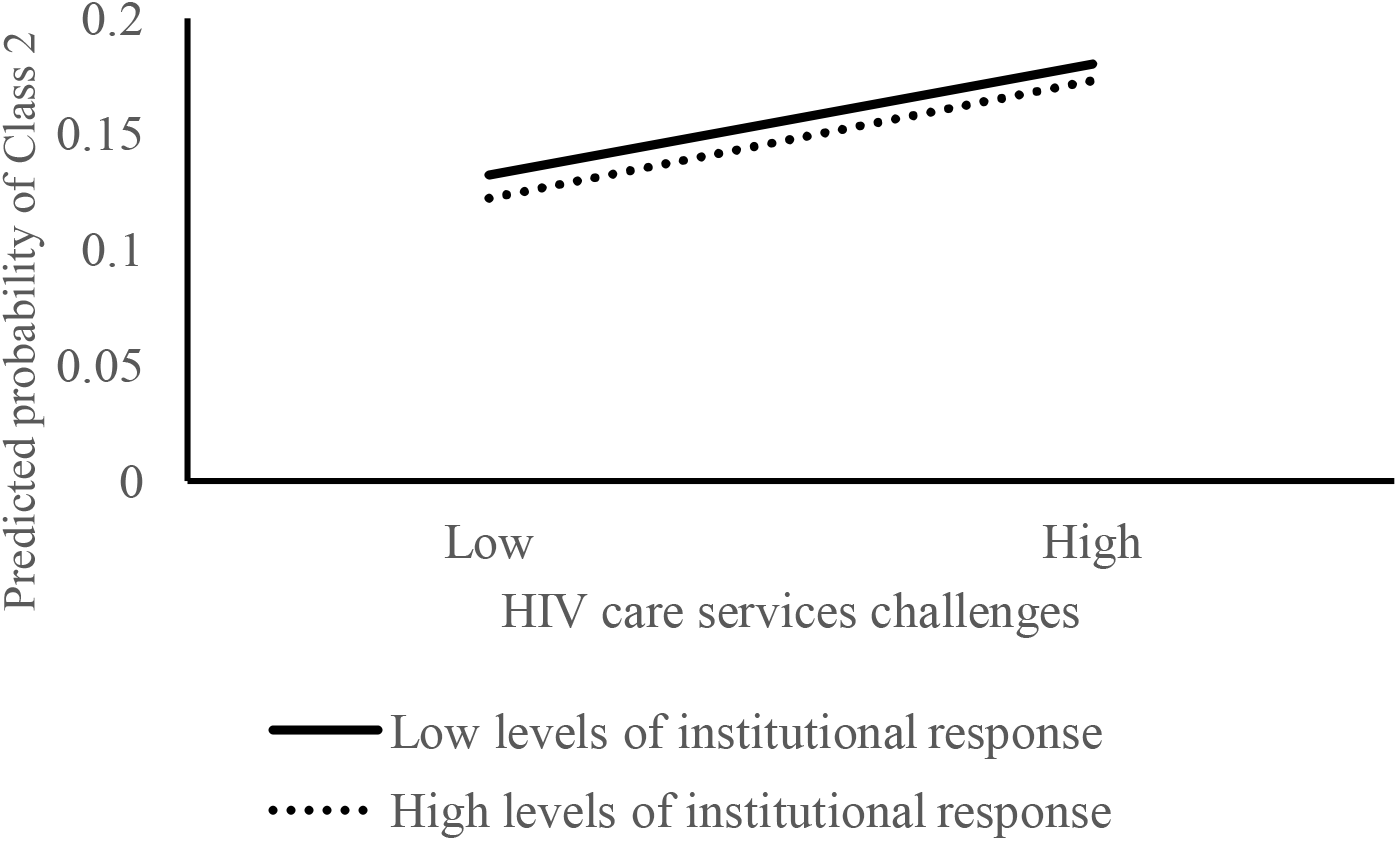
Interaction between HIV care services challenges and institutional response in the comparison between Class 2 (Moderated interruption) and Class 4 (Minor interruption) ***Note:*** *Higher and lower are defined as values above or below the mean, respectively*.

**Figure 3.**
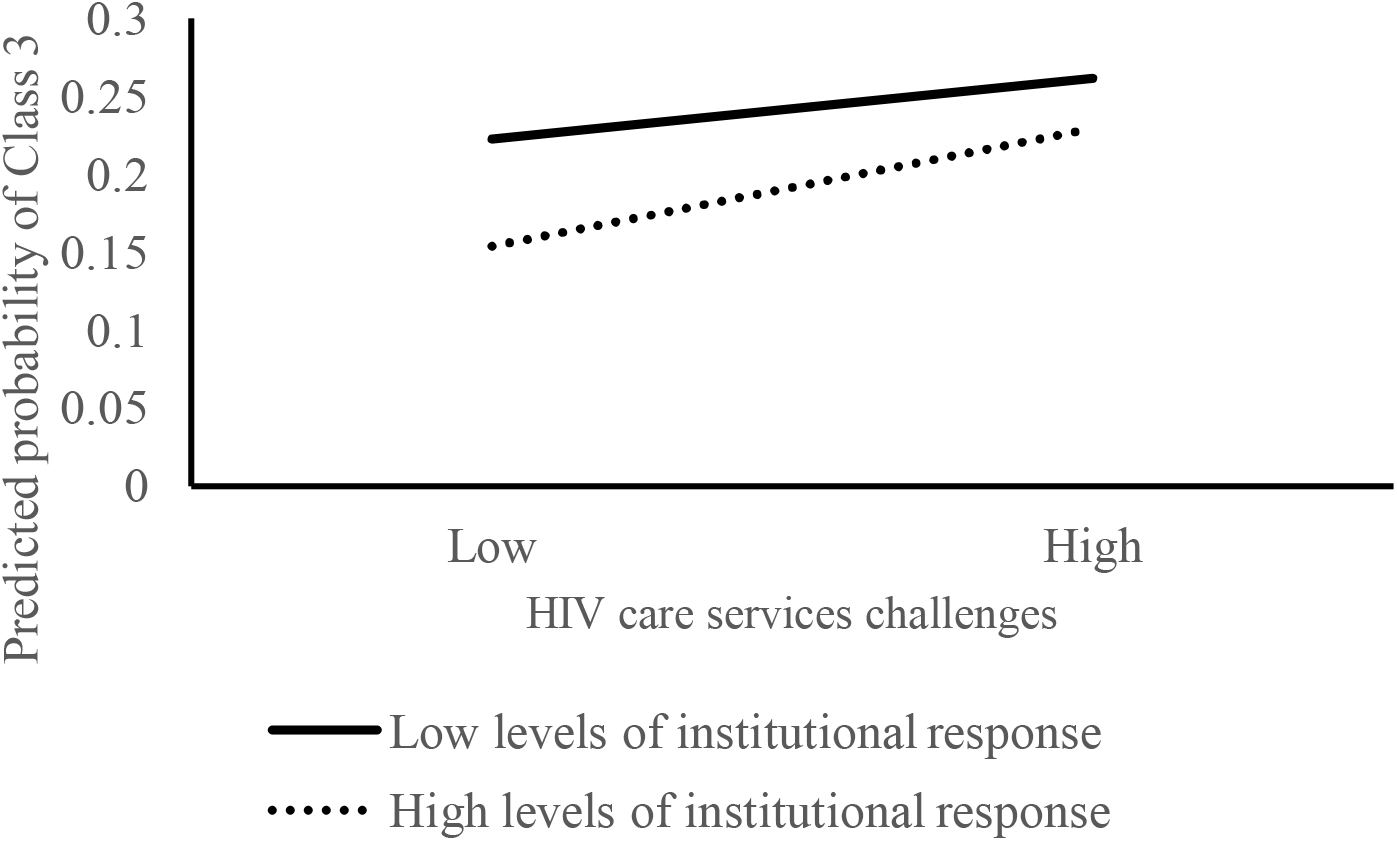
Interaction between HIV care services challenges and institutional response in the comparison between Class 3 (Almost no interruption) and Class 4 (Minor interruption) ***Note:*** *Higher and lower are defined as values above or below the mean, respectively*.

## Discussion

Using LCA, this study identified four levels of HIV service disruptions in Guangxi China during the COVID-19 pandemic, with 22.0% of healthcare providers classified as working in HIV clinics with complete interruption, 40.8% classified as working in HIV clinics with almost no interruption, and 15.4% classified as working in HIV clinics with moderate interruption. Our findings also suggested that healthcare providers who worked in county-/rural-level institutes were more likely to encounter complete interruption. Such a HIV service disruption scope is comparable with the findings in a US study (7). The HIV service delivery was more heavily disrupted in county-/rural-level institutes than province-/city-level, with the difference more evident for institutes which were completely interrupted. Institutions with more HIV service challenges are more likely to have service interruptions in all different degrees. Yet, our findings suggested a significantly negative association of institutional response only with minor interruption. The interaction effects of institutional response to HIV healthcare delivery and service challenges were only significant between the complete vs no interruption group. The findings from this study could give us a picture of HIV service interruptions in China and help guide the future preparedness of public health emergency in the context of dual epidemics.

In addition to the summarized interruption level, our results revealed particular HIV services that were likely to be disrupted during the COVID-19 pandemic. Since HIV viral suppression is critically important, thus many considerations for HIV service delivery in the context of COVID-19 focus on uninterrupted ART. Thus, it is not surprising that ART provision or refill services is rarely suspended or postponed in this study. However, around two-third health care providers reported that treatment initiation procedures were suspended or postponed for newly diagnosed HIV infection, which means that lots of newly diagnosed people could not get timely treatment after their diagnosis during the pandemic. Likewise, over half of the healthcare providers reported that the VCT services were suspended or postponed as well, followed by the outreach services. This may be because all these services require face-to-face interactions. To maintain sustainable counselling or outreach services, decentralized delivery of these services are cornerstones of the strategy, as recommended in the US (26). Decentralized delivery strategy can occur through existing or newly adopted differentiated HIV service delivery models, including community or private pharmacies, home delivery (via HIV-positive peer networks or community health centers), automated lockers, or community pickup points (e.g., post offices).

Results from the multinomial logistic regression demonstrated that the more HIV service challenges, the higher odds to have severe HIV service interruptions. To overcome the challenges of HIV service delivery in the context of COVID-19 pandemic, several measures may be beneficial for future preparation. To adapt the shortage of personnel, telehealth options, such as phone calls or other virtual options for routine or non-urgent consultations (including HIV adherence counselling), should be considered with careful scrutinize for patient privacy and confidentiality. Similar options can also be considered in place of patient support services typically offered in the community, such as peer support groups and home visits. Implementation of quarantine, social distancing, and community containment measures have reduced access to routine HIV prevention services. Adapting HIV prevention services, such as pre-exposure prophylaxis and HIV testing, may also be considered amidst the pandemic (26). HIV self-tests may be an option where traditional in-person testing services are temporarily unavailable. Even with availability of HIV self-testing kits in some areas (27), testing remains a big challenge in settings with scarce access to these kits. Thus, additional efforts are needed to augment access and to facilitate testing. For people who are already diagnosed with HIV, active patient tracking and tracing services which ensure linkage to care, and identify patients late to appointments or medication pick-ups or lost to follow up, should rely primarily on phone calls (requiring up to date contact information for all clients) before resorting to in-person tracking in communities. To support the maintenance of essential HIV service delivery, the healthcare facilities should also have a readily available sophisticated planning to deal with the dual pandemic, develop a manual or guideline for the coordination when task conflict occurs between HIV and COVID-19 service provision.

Given the effectiveness of institutional response in buffering the negative effect of HIV service challenges on complete HIV service interruption, enhancing institutional response are warranted. The HIV service interruption during the pandemic exposed the fragility of the current health system. To strengthening public health disaster risk management, resilient health system can reduce vulnerability to the public health consequences of emergent crisis (28). In the aftermath of a health crisis (e.g., COVID-19 pandemic), strong supply chain systems for essential medicines (e.g., ART supply), safe health facilities (PPE), and adequate well-trained health workers would ensure the provision of uninterrupted HIV service delivery. During this pandemic, the Chinese National Center for AIDS/STD Control and Prevention issued a notice guaranteeing a free, uninterrupted antiretroviral drug supply and released a list of ART clinics for PLWH to refill ART in China (10). A study conducted in Shenzhen, China has shared successful strategies in fighting concurrent HIV and COVID-19 pandemics (29). First, they relocated the resources of the hospital and moved the HIV clinics to an isolated clean zone to prevent nosocomial infection. The HIV care and treatment services were conducted in dedicated spaces that were physically separated from areas where COVID-19 patients were being managed. Second, the hospital minimized the number of follow-up visits by extending the medication refills for both HIV and other comorbidities, using express delivery of the medicine, online virtual platforms for counselling and consultations and medical referral, and telemedicine for PrEP and PEP services. In the non-hospital settings, facilities may consider providing services for PLWH and other chronic illnesses in the community to reduce risk of COVID-19 exposure and infection in health facilities, either using community health workers to deliver care or in makeshift clinics in the community (30). These approaches can be recommended to build the functional resilience of the healthcare system in the future public health crisis to maintain uninterrupted HIV service.

Several limitations are worth to note of this study. First, this study was cross-sectional with a lack of longitudinal follow up. Causal inference cannot be established and the results can only reflect the situation during a short period of the pandemic. Future longitudinal studies are needed to observe the evolving challenges of HIV service interruption since currently the COVID-19 pandemic was under control in China. Second, the information of HIV service challenges and service interruptions are collected based on the individual healthcare provider’s viewpoint, instead of directly measuring at the institution level. Since the number of participants were not evenly recruited from each institute, the measurement might subject to bias, which may threaten the internal validity of our findings. Third, the scales of HIV service challenges and institutional response in the study were self-developed due to the lack of established measures. These measures may not be replicated as the pandemic evolves and people’s experience change. In addition, some other institutional response to HIV healthcare delivery, such as adequate financing of emergency health service programs, strong health governance and oversight systems, might be useful to mitigate the HIV service interruptions, but not measured in our study. Given a convenience sampling approach, results in the current study may not be generalized to other Chinese regions.

As suggested by our finding, over half of institutions had a certain degree of HIV service interruptions during COVID-19 pandemic, particularly services that requires face-to-face interactions, such as VCT counselling, follow up and outreach services. HIV service challenges, such as shortage of personnel and lack of personal protection equipment, largely hinder the HIV service delivery. Institutional response could marginally buffer the negative effect of service challenges on complete HIV service interruptions comparing with no interruptions. To maintain continuity of core HIV services in face of a pandemic, build a resilient health care system with adequate preparedness is necessary, such as adapting to alternative strategies (telehealth, virtual platform, decentralized service delivery) and develop a readily available sophisticated planning (personnel arrangement, guideline or manual for coordination).

## Data Availability

Data will be available upon request from the corresponding author.

## ACKNOWLEDGMENT

Research reported in this publication was supported by the National Institute of Mental Health of National Institutes of Health under Award Number NIH/NIMHR01MH0112376 and National Science Foundation of China (NSFC) [grant number 81761128004]. The content is solely the responsibility of the authors and does not necessarily represent the official views of the National Institutes of Health.

